# Micro-Doppler Radar Identifies Movement Asymmetries After Anterior Cruciate Ligament Reconstruction

**DOI:** 10.64898/2026.04.15.26350397

**Authors:** CA Onks, C Zeng, RA Creath, BD Simone, JE Nyland, TE Murphy, LA Kishel, B Ardat, VA Venezia, AM Wiggins, BR Shaffer, RM Narayanan

## Abstract

**Background:** Patients who have undergone Anterior Cruciate Ligament Reconstruction (ACLR) have a 6-24% chance of either re-tearing or having subsequent knee surgery. To date there have been no practical validated risk prediction models that can be easily implemented into clinical workflow for re-injury risk. Micro-Doppler radar (MDR) provides a promising solution.

**Objective:** The purpose of this study was to investigate the predictive ability of MDR to identify persons with a previous ACLR relative to an age and sex matched healthy control.

**Methods:** ACLR patients (n=81) and controls (n=100) performed drop box jump, sit to stand (STS), and walking trials as MDR signatures were collected. A 1D Convolutional Neural Network was developed to evaluate each activity individually followed by the development of a fusion model validation using all three activities.

**Results:** The STS model individually achieved the highest overall accuracy of 82.3%, with a sensitivity of 71.6% and specificity of 91.0%. The fusion model using all activities achieved a peak overall accuracy to detect ACLR of 86.2%, 80.3% sensitivity, and 91% specificity.

**Conclusions:** Currently, there is no clinically validated, efficient approach to objectively evaluate human motion at the point of care. When coupled with machine learning, MDR accurately differentiates ACLR from control groups by identifying complex biomechanical asymmetries, with classification performance comparable to or exceeding that of motion capture. Future research is needed to determine if MDR can be used in conjunction with risk prediction modeling.

**Key points:** Micro-Doppler radar provides a promising new solution to identify important human motion asymmetries in clinical settings. Here we evaluated a group of patients who have a history of Anterior Cruciate Ligament reconstruction versus a control group. Simple movements performed in the presence of the micro-Doppler radar system were used to identify the 2 groups with accuracy comparable or superior to motion capture systems.

## 1 Introduction

### 1.1 The Burden of Anterior Cruciate Ligament Injury

Anterior Cruciate Ligament injuries (ACLI) are common knee injuries in the United States and comprise more than 50% of knee injuries.[1] It is estimated that more than 200,000 Americans are affected annually at a cost of $ 7 billion between indirect and direct costs [1]. These injuries primarily occur in those physically active, including athletes and military service members under the age of 30, with a peak incidence of 226.7 per 100,000 in the ages of 14 to 18 years [2], [3], [4].

Surgical reconstruction following ACLI continues to be the recommended treatment for athletes [1]. Unfortunately, patients who have undergone Anterior Cruciate Ligament Reconstruction (ACLR) have a 6-24% chance of either re-tearing or having a subsequent knee surgery on either side after successful completion of surgery and post-surgical rehabilitation [5]. In addition, one third of these patients will experience post-operative osteoarthritis 10 years after ACLR, despite undergoing anatomic surgical repair [6]. This leads to poor athletic performance, discontinuation of sports, and reduction of physical activity levels, all of which negatively influence health-related quality of life [3].

Multiple modifiable and non-modifiable risk factors have been identified for ACLI, but there is a need to clarify how these risk factors can help practitioners develop prevention guidelines [7], [8]. Injury prevention programs have been validated to decrease the incidence of ACLI [9–11]. Unfortunately, studies on implementing these programs report difficulties with poor adoption, compliance, and maintenance of injury prevention interventions [12]. One of the challenges is applying these programs to the general population. These programs take time and extensive expertise to administer, which can be prohibitive. In other medical settings, prevention efforts are typically risk-based, seeking to identify those most at risk so that limited preventative resources can be directed to the highest-risk population. Examples would include primary prevention for infectious diseases or cancers through vaccination programs or cancer screenings [13, 14]. This has not been the approach to mitigation or prevention of ACLI, as risk prediction models are lacking in the routine care of those at risk for ACLI.

Biomechanical abnormalities, which include differences in movement, force development, extremity loading asymmetries, and lower jump heights, have been linked to ACLR [15–19]. These asymmetries are potential targets to understand the risk of re-injury, and have been targets for injury prevention programs, but clinical assessment of these abnormalities has proven challenging. To date, there have been no practical, validated risk prediction models that can be easily implemented into clinical workflow or deployed for military use [20–22]. A recent scoping review found nearly 2,000 publications on research looking to identify ACLI risk [23]. The results were disappointing as there were no screening tests that could identify injured versus uninjured groups [23]. In another review, it was reported that 79% of current prediction models were rated as high risk for bias, poorly developed, and not applicable for further model validation by other researchers [24]. For all of these reasons, it is imperative that we continue to seek alternative approaches to identify those most at risk for ACLI or re-injury.

### 1.2 Micro-Doppler radar

The use of Micro-Doppler Radar (MDR) is a novel approach for evaluating human mechanics [25–28]. Radar, in its simplest form, is a device consisting of a radio transmitter, a radio receiver, and two antennas (Figure 1). Typically, radars are designed to detect the position of a target and infer the speed of a moving target using the Doppler principle [29]. When a radar target’s bulk motion produces a constant Doppler frequency shift, additional micro-motions such as vibrations or rotations of structures on the target will create extra Doppler modulations in the returned signal, known as the micro-Doppler effect [30]. Human walking involves multiple coordinated movements as follows: the body mainly translates forward with slight rocking and head motion, while the arms and legs swing back and forth, producing strong micro-motions from partial rotations around joints. As a result, human movements produce unique micro-Doppler signatures (MDS) represented in spectrograms seen in Figure 2, which are changes in frequency over time produced by the movement that occurs in front of the MDR. In particular, MDS can identify human gait patterns and other activities by discerning both gross and fine movements, even if the latter are so subtle that they are indistinguishable to direct observation by a skilled clinician [31–33]. Standard human biomechanical analyses, such as motion capture systems, have not previously been able to capture movements at this resolution [34]. Previous work has shown that MDR coupled with machine learning can identify subtle biomechanical differences that can identify pathologic gait patterns, those at risk of musculoskeletal injury, and those at risk for falls [25, 35–38].

**Fig 1.**
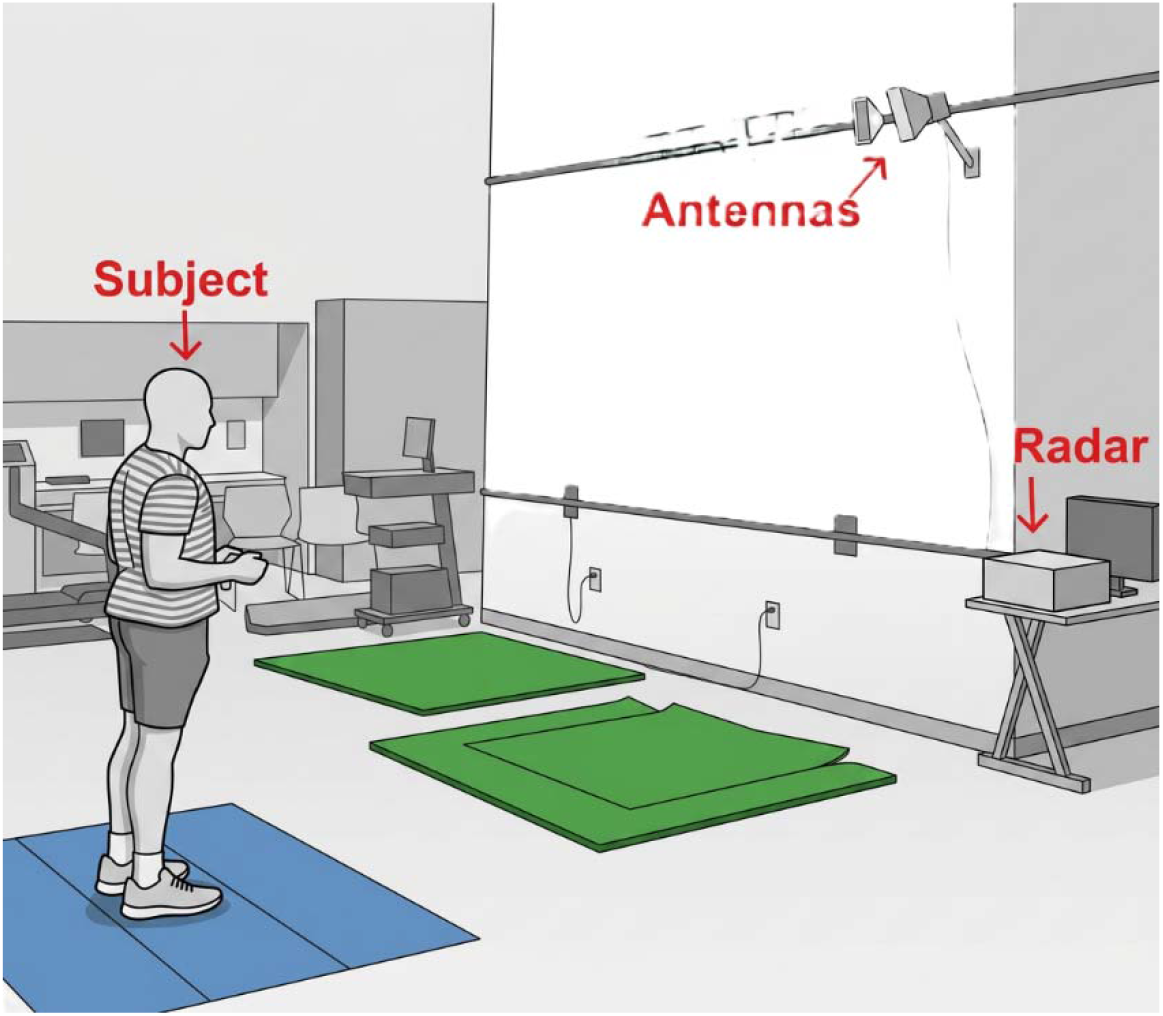
Study protocol set up demonstrating the relationship of the subject to the antenna and micro-Doppler radar

**Figure 2:**
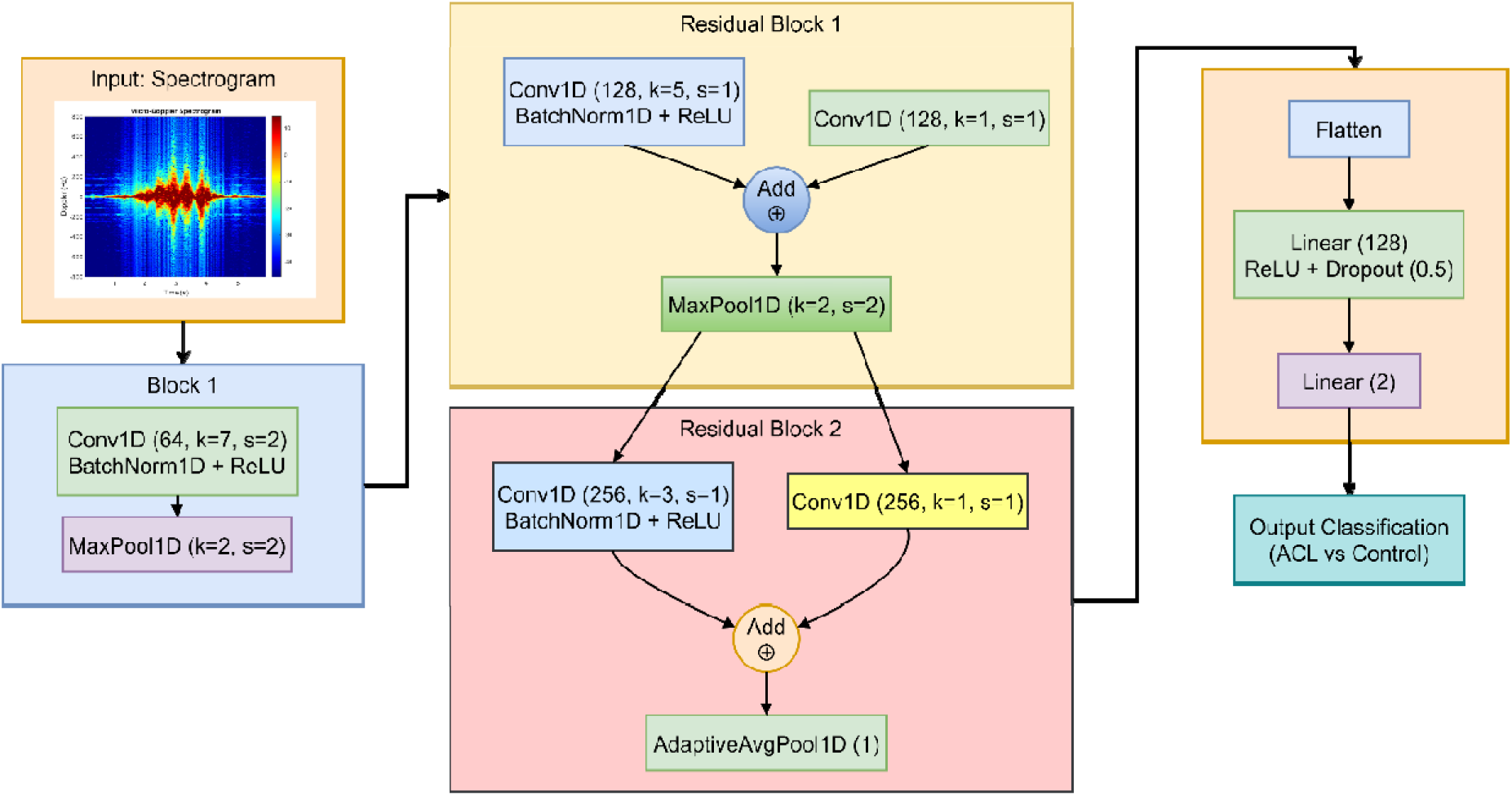
Deep Learning network ^*^Abbreviations; Conv1D – One-Dimensional Convolutional Layer, K – Kernel Size, S – Stride, BatchNorm1D – One-Dimensional Batch Normalization, ReLU – Rectified Linear Unit, MaxPool – Max Pooling Layer, AdaptiveAvgPool1D – One-Dimensional Adaptive Average Pooling Layer

Due to the burden of ACLI, there is a need for practical clinical approaches to assess ACLI risk and risk or re-injury. The purpose of this case-control study was to evaluate the predictive ability of MDR to differentiate between those with a prior ACLR and healthy controls matched by age and sex. We hypothesize that MDR has the potential to accurately differentiate subjects with ACLR from uninjured matched controls.

## 2 Methods

### 2.1 Study Design

We completed an IRB-approved (STUDY00020118) case control study between February 2023 and May 2025 to compare adults with previous ACLR to age and sex matched healthy adult controls. This study is part of a larger research initiative described in the protocol publication [39]. Subjects with a history of ACLR were identified by review of the electronic medical record from a large regional health system. Recruitment letters were sent to eligible participants; if no response was received, a phone call was made to the potential participant. The control group was recruited from the surrounding community through advertisement flyers in the same medical system that the ACLR group was recruited from, and at a local college campus where the data collection took place. Our research coordinator contacted the potential subjects, who were first screened for eligibility, and verbal consent was obtained while remaining compliant with all Health Insurance Portability and Accountability Act requirements. Once enrolled, the subjects were then scheduled for data collection.

### 2.2 Participants

Inclusion criteria for the ACLR group included: 1) age 18-40 years, 2) history of ACLR in the past 9 months to 6 years, and 3) surgeon’s clearance to return to normal activities. Individuals were excluded from our study for: 1) pregnancy, 2) institutionalization, 3) history of cerebral vascular accident, 4) inability to complete informed consent, 5) inability to perform protocol activities, 6) history of knee or hip replacement, 7) inability to walk or jump without a limp, and 8) current neuromuscular disease. The inclusion criteria for the healthy control group were: 1) age 18-40 years, and 2) never having had a lower extremity surgery. The exclusion criteria were identical to the ACLR cohort.

### 2.3 Experimental Protocol and Measurement

Following informed consent, participants were asked to complete a demographics survey available through REDCap, a secure data information system commonly used in research studies [40]. Collected socio-demographics included: 1) age, 2) sex, 3) body mass index, and 4) race.

We then collected radar MDS on the selected participants in a state-of-the-art human movement laboratory. Participants were asked to perform three functional activities selected to cover a range of performance abilities: 1) two-leg drop box jumps (DBJ), 2) standing from a seated position (STS), and 3) overground walking (WLK).

Several researchers have used DBJ to assess ACLI risk [41–46]. Motion capture studies have identified variables such as valgus of the knee during vertical landing propulsion phases immediately prior to landing and inter-knee distance during the compression and propulsion phases to be risk factors for ACLI [43]. Similarly, motion capture studies have identified biomechanical differences following ACLR versus nonsurgical control groups [18, 47]. For this reason, DBJ was chosen to evaluate whether MDR could also identify subtle changes from performing this activity. Subjects were asked to stand on a heavy 30-cm high box with their hands resting on their waist. At the subject’s discretion, they would step off the box such that their fall distance is equal to the box height and land with both feet simultaneously on the floor. Upon landing, they would immediately and without delay execute a vertical jump landing in place (without horizontal translation). The trial ended when the subject returned to the upright standing position following their jump. Subjects performed five trials with 30-60 seconds of rest in between trials.

Similarly, STS has been studied specifically following ACLR, in which those injured have been shown to shift their weight forward relative to foot placement on the side of the ACLI [15, 17, 48]. These biomechanical differences were the basis for using STS. Subjects began by sitting on a height-adjustable seat. The seat was flat and did not have arms. Seat height was adjusted so that the angle between the thigh and shank was 90 degrees when seated with feet flat on the floor. Initial trunk position was vertical with the arms crossed across the chest. At the subject’s discretion, they would stand up as quickly as possible, achieving a final stationary vertical standing position. The trial would end after the subject maintained the final stationary standing position for five seconds. Subjects performed five trials with 30-60 seconds of rest in between trials.

Finally, WLK was selected as a common everyday activity that has demonstrated variable motion capture findings for the identification of Anterior Cruciate Ligament (ACL)-related deficiencies. Kinematic gait measures have been identified as clinically significant to identify the ACLI participants relative to the uninjured participants, but this was not statistically significant [49]. In a more recent study, authors found statistically significant ACL-related deficiencies for treadmill walking on a surface with 10-degree upward, level, and 10-degree downward slopes for peak knee flexion and peak knee displacement [50]. Additional support for the use of WLK comes from the MDR literature, where preliminary gait studies have demonstrated successful differentiation of gait patterns [35, 51, 52]. Due to the varying outcomes with motion capture-based research and promising MDR gait research, we chose WLK to evaluate whether MDR would clearly differentiate the ACLR group versus control. Subjects were asked to perform three overground walking trials on a 10-meter level surface walking towards the antenna along the central axis of the radar at their preferred gait velocity. Gait initiation and termination were included in the analysis.

#### 2.3.1 Radar Parameters and Set-Up

The components of the radar were purchased, assembled, and calibrated prior to data collection. The MDR system used in this study was a custom-designed, portable continuous-wave (CW) radar operating in a quasi-monostatic configuration at a 10-GHz carrier frequency in the X-band. This frequency was chosen to balance component availability, cost, and a wavelength suitable for capturing subtle biomechanical variations in human motion. The hardware specifications were completed based on previously published parameters [34, 39, 53]. The radar was placed outside of the data collection area. The antennas were mounted at a height of 3 meters and 3 meters from the participant at 40 degrees down from horizontal, as seen in Figure 1. All participants performed the protocol as described.

#### 2.3.2 Data Processing, Activity Classifiers, and Validation

##### Signal Processing and Spectrogram Generation

Raw in-phase and quadrature radar signals were initially captured at a sampling rate of 44.1 kHz using Audacity software. Data processing was subsequently performed using MATLAB (Signal Processing Toolbox, R2023b, MathWorks, Natick, MA). Signals were then down sampled to 2 kHz due to human gait features of interest falling below 1 kHz. Time-domain signals for DBJ, STS, and WLK tasks were transformed into time-frequency spectrograms using a Short-Time Fourier Transform (STFT). We employed a 128 ms (256 samples) Hamming window with 75% overlap and a 1024-point Fast Fourier Transform. These parameters were selected to provide enhanced frequency resolution necessary for distinguishing subtle micro-Doppler features. The resulting magnitude spectra were log-transformed to compress the dynamic range and Z-score normalized to ensure feature consistency. Finally, all spectrograms were standardized to a dimension of 1024 × 512 (frequency bins × time steps) via cubic interpolation to serve as inputs for the neural network.

##### Deep Learning Activity Classification

We used a 1D Convolutional Neural Network (CNN1D) to evaluate the spectrograms for each activity represented in Figure 2. This architecture treats the 1024 frequency bins as input channels to extract hierarchical temporal features. Model training utilized the AdamW optimizer combined with a Cosine Annealing learning rate scheduler to ensure optimal convergence. To address potential class imbalances and improve generalization, we employed a class-weighted cross-entropy loss function with label smoothing. An early stopping mechanism was implemented to prevent overfitting by monitoring validation loss during training. This final model for each of the activities was then ready for testing.

##### Fusion Model Validation Framework and Statistical Metrics

To maximize classification performance, we implemented a stacking ensemble framework. The independent CNN base models trained for each activity (STS, DBJ, WLK) were fused using a meta-classifier. We evaluated four distinct meta-modeling algorithms to determine the optimal fusion strategy: Logistic Regression, Support Vector Machine with Radial Basis Function kernel (SVM-RBF), Gradient Boosting, and Random Forest.

To rigorously evaluate model generalizability and prevent data leakage, a hierarchical cross-validation strategy was employed. For our base model activities, we utilized a 20-fold Stratified Group K-Fold cross-validation. The “Group” constraint ensured that all trials from a single participant were assigned exclusively to either the training or testing set within any given fold, guaranteeing participant-level independence. For the fusion models, we used separate 5-fold Stratified Group K-Fold cross-validation on the aggregated probability vectors to validate the meta-classifiers.

In a Threshold-Aware Cascade analysis, we sought to further refine and optimize the performance of the fusion model using a Threshold-Aware Cascade strategy. In this analysis, a decision rule was created in which the activity demonstrating superior performance would act as the primary decision as the “expert.” If the model’s confidence score fell within a high-certainty range, its prediction was accepted directly. For uncertain cases, the decision was deferred to the ensemble meta-classifier (Logistic Regression). The optimal confidence threshold range ([0.0, 0.60]) was determined through a rigorous grid search optimization involving 121 potential combinations to maximize validation accuracy.

## 3 Results

### 3.1 Sample Inclusion and Demographics

Recruitment into the ACLR group was targeted via an electronic medical record query, which identified 523 patients aged 17 to 40 years who underwent ACLR surgery between March 2018 and March 2024. The list of patients included those who had undergone ACLR surgery at age 17, but potential participants were not approached for enrollment until they turned 18. We ultimately mailed 416 recruitment packets that contained an introduction letter, study summary, response form for participants to indicate their interest in the study or request no further contact, and a self-addressed, postage-paid envelope for easy return of the response form to the study team. Of the 416 potentially eligible patients, 24 could not be reached (e.g., the recruitment letter was returned as undeliverable, phone numbers were incorrect or disconnected), and 13 returned the response form requesting no further contact about the study. If a participant didn’t return the response form requesting no further contact, the study team called them 2-3 weeks later.

In total, 38 ACLR patients were excluded. Common causes of ineligibility were non-ACL surgeries within the previous 6 months, having an ACLR greater than 6 years from or less than 9 months prior to enrollment, inability to attend the study visit, polytraumatic injury, and pregnancy. A total of 201 potential participants did not respond, and 72 responded, indicating that they were not interested in participating.

Additional ACLR and CTRL subjects were also recruited via flyers hung throughout the health system and affiliated college of medicine campus, local physical therapy offices, and local college campuses. A research recruitment database (*Studyfinder*) website was also utilized.

A total of 263 potential healthy participants responded to recruitment advertisements for enrollment into the CTRL group. Of these, 33 were deemed ineligible due to age, inability to attend the study visit, surgery in the previous 6 months, or pregnancy. Ultimately, 81 ACLR subjects and 100 CTRL subjects were successfully enrolled. The two groups were age and sex-matched. Table 1 provides the demographics for the subjects who were enrolled and ultimately completed the study protocol.

**Table 1.**
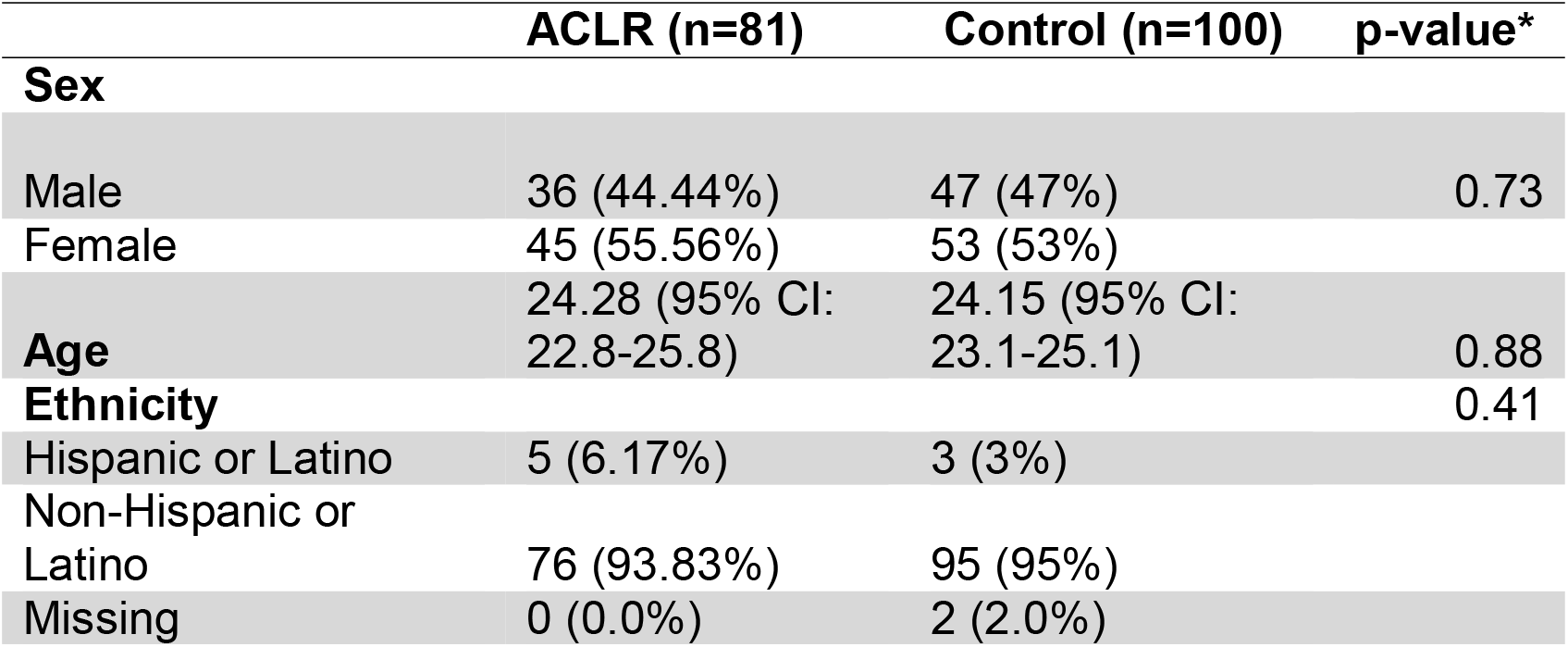
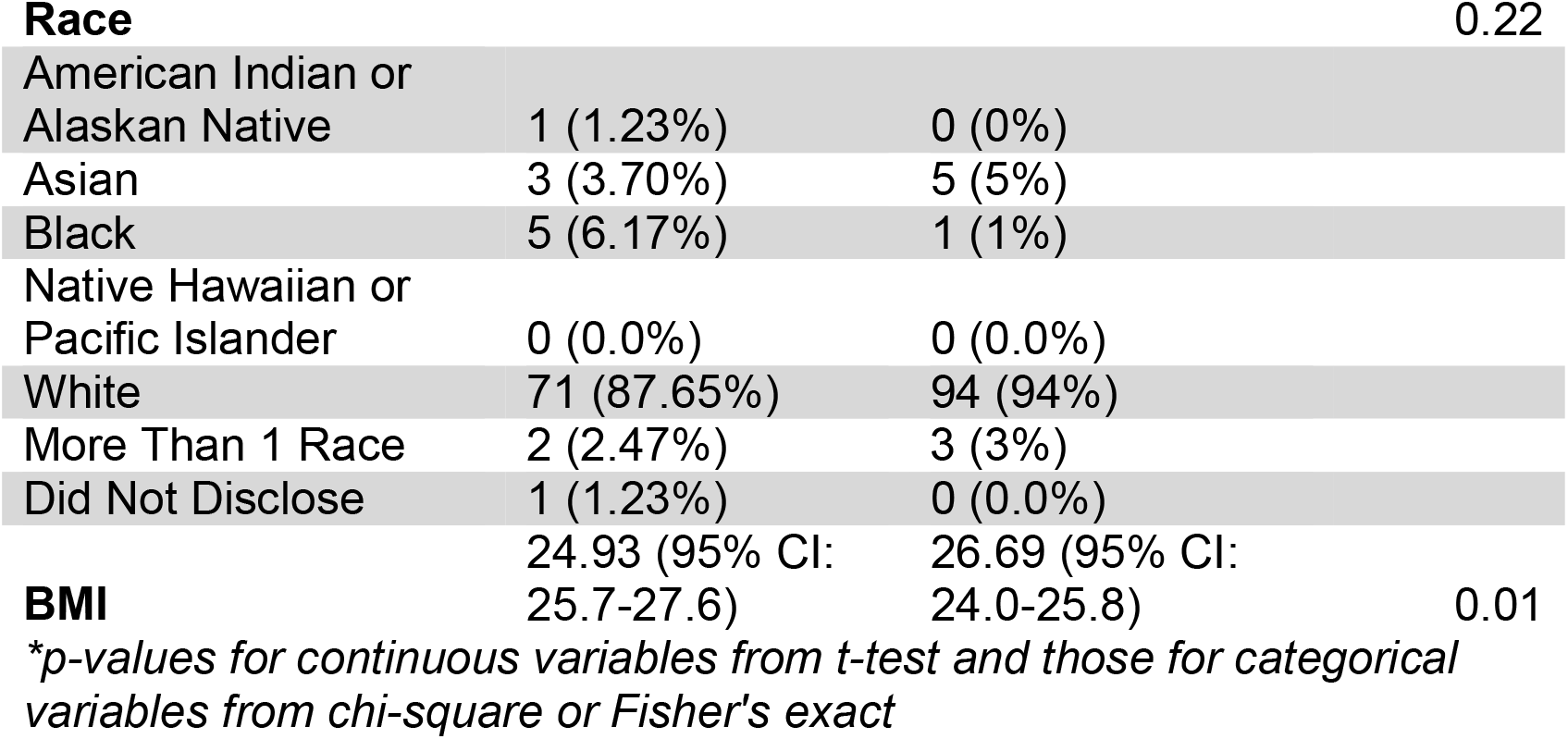
Demographics and clinical characteristics. *Abbreviations; BMI- Body Mass Index, ACLR - Anterior Cruciate Ligament Reconstruction

### Spectrogram Generation

The study protocol was completed by enrolled participants, producing three data sets for analysis. There were 905 DBJ trials, 905 STS trials, and 544 WLK trials. The data from each trial were processed, yielding a spectrogram for each individual trial. Figure 3 shows a single representative spectrogram for each activity in the ACLR and CTRL

**Fig 3.**
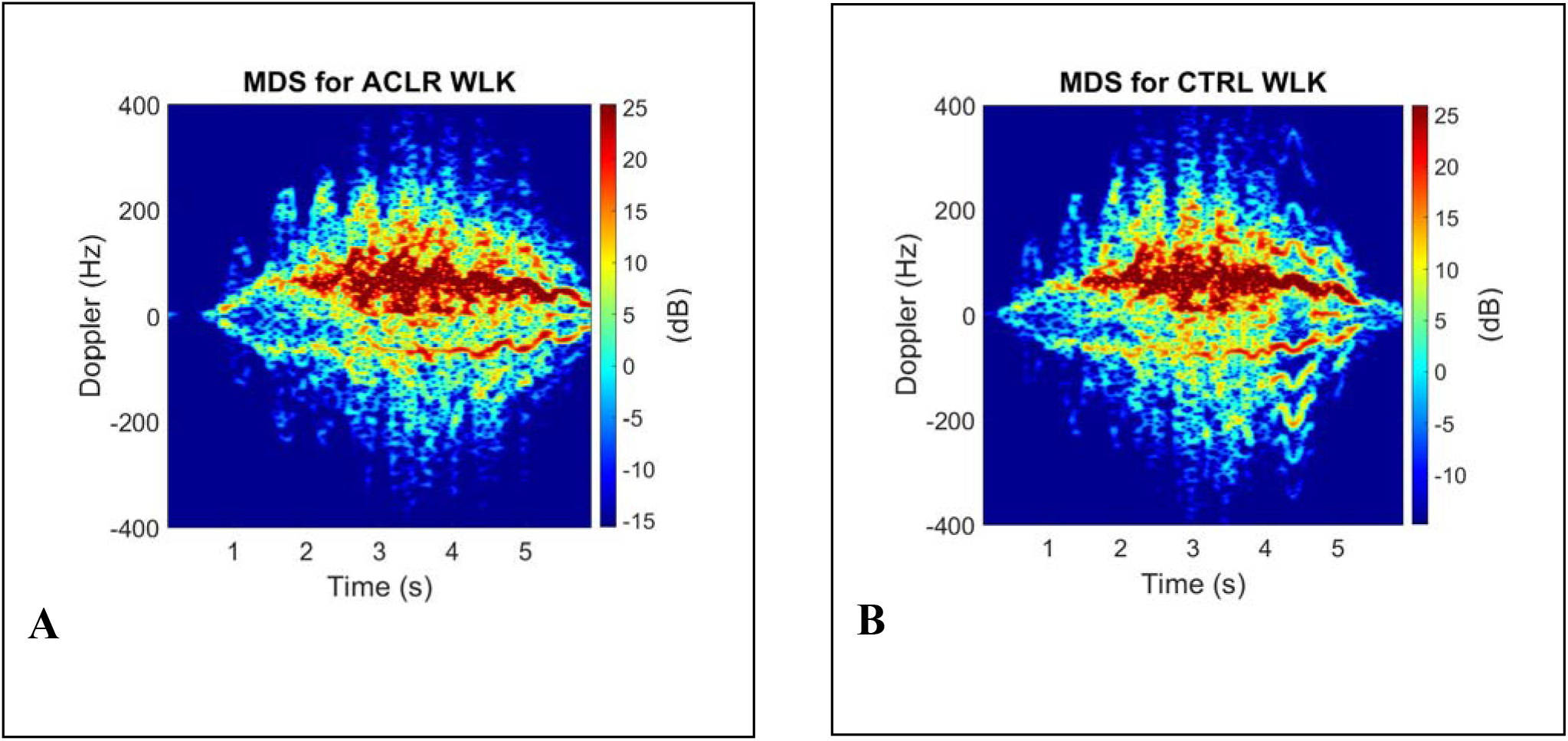
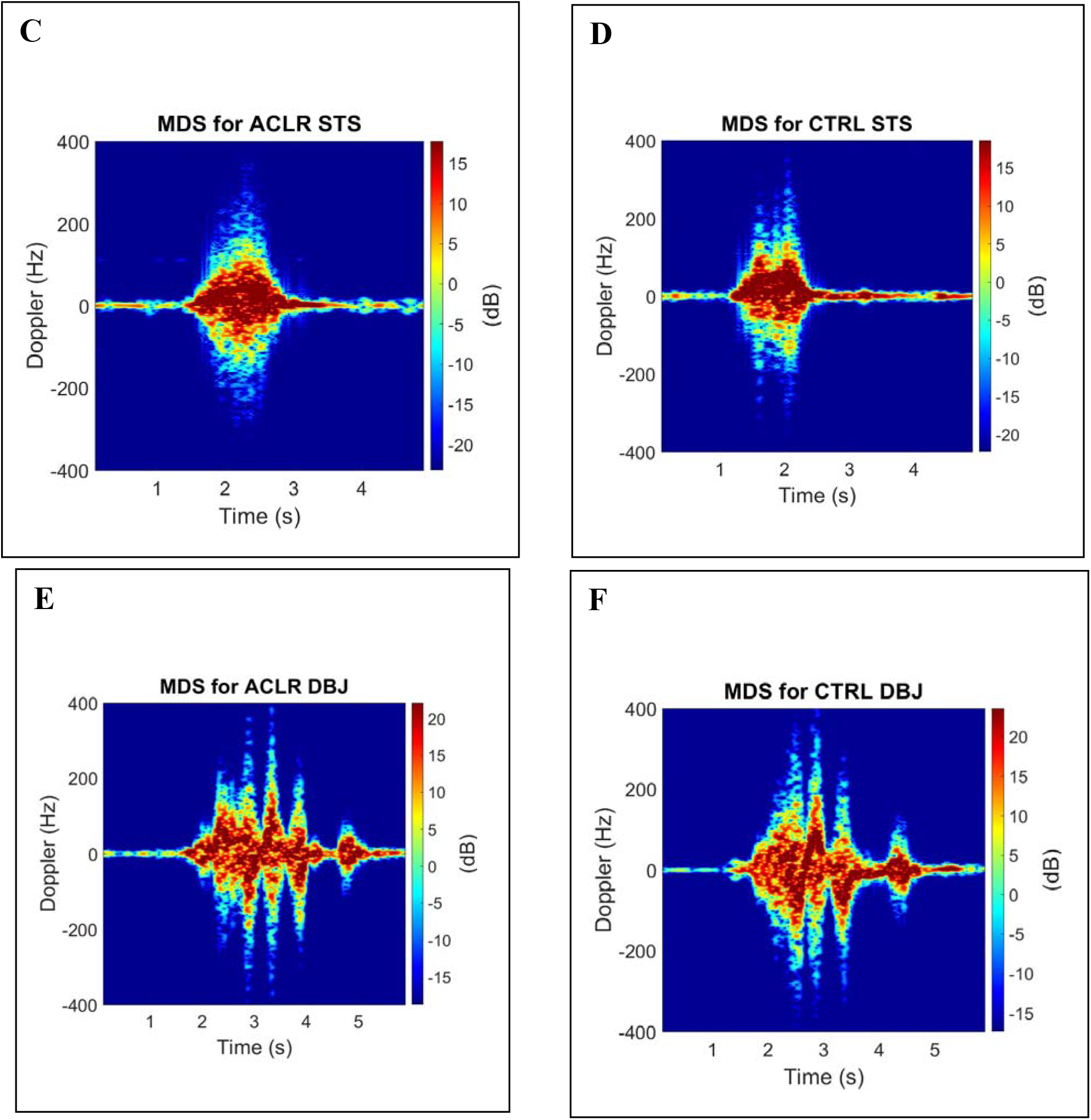
Processed **s**pectrograms for individual activities developed for deep learning (**A)** ACLR WLK (**B**) CTRLWLK (**C**) ACLR STS (**D**) CTRL STS (**E**) ACLR DBJ (**F)** CTRL DBJ. ^*^Abbreviations; MDS - Micro-Doppler Spectrogram, ACLR – Anterior Cruciate Ligament Reconstruction, CTRL – Control Group, WLK – Overground Walking, STS – Standing from a seated position, DBJ – Two-leg Drop Box Jump

### 3.3 Activity Classification

Deep learning models were applied to the spectrograms, and the individual functional activities listed in Table 2 were trained and tested. The STS model achieved the highest overall accuracy of 82.3%, with a sensitivity of 71.6% and a high specificity of 91.0% to identify ACLR. The DBJ model yielded an overall accuracy of 81.8% (Sensitivity: 75.3%, Specificity: 87.0% for ACLR). Finally, the WLK model produced an overall accuracy of 81.3% (Sensitivity: 75.6%, Specificity: 86.0% for ACLR).

**Table 2:** Performance of deep learning models for identification of the ACLR versus CTRL completed for each individual functional activity. *Abbreviations; STS – Standing from a seated position, DBJ – Two-leg Drop Box Jump, WLK – Overground Walking

| Activity | Overall Accuracy (%) | Sensitivity (%) | Specificity (%) |
| --- | --- | --- | --- |
| STS | 82.3 | 71.6 | 91.0 |
| DBJ | 81.8 | 75.3 | 87.0 |
| WLK | 81.3 | 75.6 | 86.0 |

### 3.4 Fusion Model Performance

The comparative analysis of fusion strategies is presented in Table 3. The fusion meta-models were then evaluated separately using 5-fold cross-validation on the aggregated participant-level probability outputs. Standard stacking approach accuracies, such as Random Forest (77.9) and Gradient Boosting (77.9%), underperformed compared to individual models with SVM-RBF (81.8%) and Logistic Regression (81.2%), improving performance over individual models. The Threshold-Aware Cascade strategy yielded superior diagnostic performance. By leveraging the high specificity of the STS task and correcting its uncertainties with the ensemble committee, this method achieved a peak overall accuracy of 86.2%, with 80% sensitivity to detect ACLR, and 91% specificity to identify healthy controls. Figure 4 shows the final confusion matrix (CM) classification decisions for the ACLR and control groups in each of the fusion strategy models. This was generated from the aggregated 5-fold Stratified Group K-Fold cross-validation to provide highly robust estimate of the model’s performance.

**Table 3:** Fusion Model Strategy Performance for the identification of the ACLR versus CTRL groups. *Abbreviations; ACLR – Anterior Cruciate Ligament Reconstruction, CTRL – Control Group, SVM-RBF – Support Vector Machine with Radial Basis Function

| Fusion Strategy | Overall Accuracy | Sensitivity (ACLR) | Specificity (CTRL) | F1 Score (ACLR / CTRL) |
| --- | --- | --- | --- | --- |
| Threshold-Aware Cascade | 86.2% | 0.80 | 0.91 | 0.84 / 0.88 |
| Logistic Regression | 81.2% | 0.74 | 0.87 | 0.78 / 0.84 |
| SVM-RBF | 81.8% | 0.79 | 0.84 | 0.80 / 0.84 |

Table 3: Fusion Model Strategy Performance for the identification of the ACLR versus CTRL groups
| **Fusion Strategy** | **Overall Accuracy** | **Sensitivity (ACLR)** | **Specificity (CTRL)** | **F1 Score (ACLR / CTRL)** |
| --- | --- | --- | --- | --- |
| **Gradient Boosting** | 77.90% | 0.77 | 0.79 | 0.76 / 0.80 |
| **Random Forest** | 77.90% | 0.77 | 0.79 | 0.76 / 0.80 |

**Figure 4:**
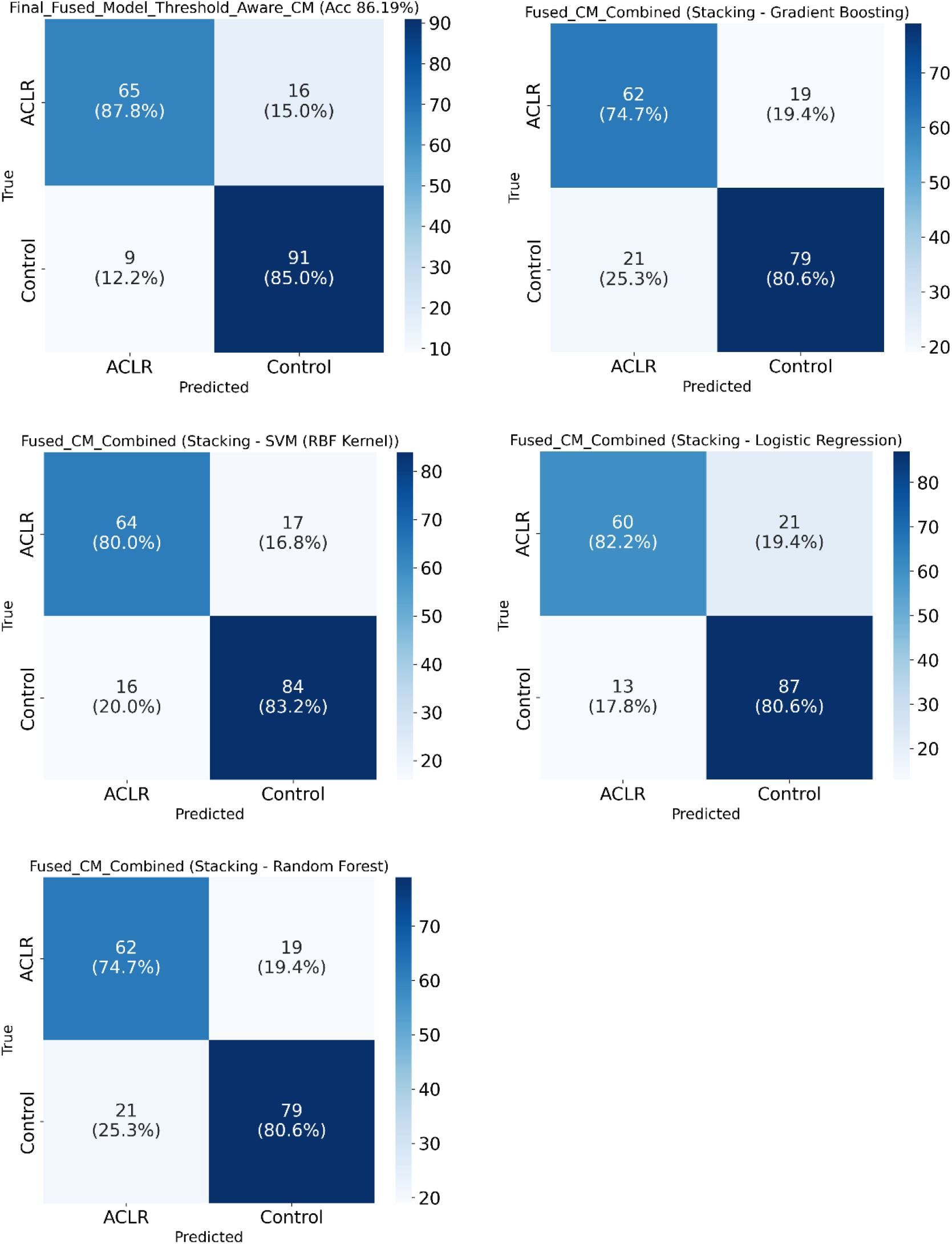
Confusion Matrix providing final classification performance ^*^Abbreviations; CM - Confusion Matrix, ACLR – Anterior Cruciate Ligament Reconstruction, SVM-RBF - Support Vector Machine with Radial Basis Function

The Threshold-Aware Cascade analysis of the decision path revealed that 94 participants (52%) were classified directly by the STS “expert” model with high confidence, while the remaining 87 participants (48%) required the “committee” (ensemble model) to resolve uncertainty, demonstrating the efficiency of this hierarchical approach.

## 4 Discussion

We have demonstrated that MDR-based algorithms were able to successfully determine that participants had a history of ACLR by observing individually either DBJ, STS, or WLK with an accuracy of over 81%. Significantly, the Threshold-Aware Cascade fusion model further improved diagnostic performance to a peak accuracy of 86.2% by fusing the three activities. This is the first time MDR has been used to observe simple functional activities analyzed using machine learning to identify unique micro-motions in the radar signal that would differentiate specific patient populations, such as those with a history of ACLR. This is a particularly important finding as MDR is a practical, portable, low-cost, and reliable technology that can identify ACLR with accuracy comparable to previous motion capture studies [45].

### 4.1 ACLR versus CTRL Analysis

For the primary analysis of individual activities, we have demonstrated that MDR fused with machine learning can differentiate imperceptible biomechanical differences in spectrograms between the ACLR and CTRL groups. This was accomplished by performing STS, DBJ, and WLK activities in the presence of MDR. Each activity performed individually was able to differentiate the ACLR group from CTRL with high accuracy. STS had the best accuracy at 82.32%, and its dominant role in the fusion model underscores its clinical value, demonstrating that an activity as simple as standing from a chair may provide significant clinical insight when observed with MDR. This is important to demonstrate that MDR can identify underlying biomechanical differences in patient populations, such as ACLR in the MDS. Additionally, our WLK MDS model resulted in 76% sensitivity and 86% specificity to identify the ACLR versus control, where previous motion capture data have had mixed results for using WLK [49, 50]. These findings confirm that MDR can detect biomechanical asymmetries within specific patient populations that are comparable and potentially an improvement to sophisticated motion capture labs especially when considering the practical benefits of MDR.

Patients who undergo ACLR go on to re-tear their ACL or have a second surgery 6-24% of the time, which was the basis for using this patient population, in addition to known leg asymmetries that persist long after repair and rehabilitation. [5, 45, 48]. It is felt that biomechanical alterations at least in part are linked to repeat injury following ACLI, as shown by previous motion capture data demonstrating altered strength and alignment after ACLR and return to athletics [19, 54, 55]. MDR has demonstrated high accuracy in detecting differences between the ACLR and CTRL groups. Unlike single-activity models common in motion capture settings, our fusion strategy leverages the STS “expertise” as the first line of classification, offering a highly efficient alternative to traditional motion capture systems while maintaining superior accuracy. Importantly, MDR could serve as an alternative approach to identifying persisting asymmetries following ACLR.

While individual tasks performed well, the Threshold-Aware Cascade model achieved a superior accuracy of 86.2%. This strategy maximizes clinical efficiency: the model first attempts to classify the patient using only the STS task; if the confidence is below a certain threshold, it then incorporates WLK and DBJ data. This ‘expert-first’ approach mimics clinical triaging, where simple tests are used initially, and more complex assessments are reserved for ambiguous cases.

Finally, we attempted to understand how our model best learned from the fusion model to better understand the black box phenomenon described in machine learning models. In this analysis, the results indicated that the STS task provided the highest discriminative power. This is helpful as this hierarchy justifies the design of our Threshold-Aware Cascade model, where the STS task acts as the primary “expert” classifier, which was chosen because it had the highest overall accuracy when the CNN model was applied only to this task. Evidence from motion capture studies indicates STS exposes biomechanical deficits following ACLR, lending support to our findings where these deficits manifest as distinguishable MDS between ACLR and CTRL.

### 4.2 Clinical Application

Here, we have demonstrated that MDR coupled with machine learning is able to identify ACLR versus a control group with great accuracy. This supports our hypothesis that MDR can identify subtle asymmetries associated with human movements, such as those resulting from ACLR. Radar was initially used primarily for military applications such as target detection and terrain imaging. However, over the past 40 years, it has been applied to civilian use, such as resource monitoring, nondestructive testing, and automotive sensors. It is only in recent years that attention has been turned to human-based monitoring to identify when falls may occur or who is at risk of falling [56–58].

MDR technology has been shown to be reliable and cost-effective over many years of use. We were able to complete our study with one radar unit collecting over 180 hours of data with continuous monitoring, demonstrating its endurance and ease of use. Once MDR algorithms are trained to identify patient populations, such as ACLR, these can be used clinically. In this case, we could use the ACLR MDR algorithm in post-operative management to determine whether asymmetries persist after ACLR. Once ACLR patients are identified as having persistent deficits, targeted and validated neuromuscular training programs can be implemented for those who continue to exhibit asymmetries, thereby decreasing injury risk [9, 44].

ACLR was used here as a model and proxy for broader clinical applications of this technology across Orthopaedics, Primary Care, Sports Medicine, Rehabilitation, or any musculoskeletal disease management. There are currently no clinically efficient movement-related screens readily used in practice that can differentiate at-risk groups with the accuracy we have demonstrated. MDR coupled with machine learning represents an exciting new technology with the potential to evaluate musculoskeletal injury (MSKI) risk in ways that have not been clinically feasible previously. MDR technology provides an ideal solution as it is small, portable, inexpensive, deidentified, not reliant on light, and has been shown to distinguish very small differences between movement patterns in our preliminary work [34, 53]. We have now demonstrated that MDR-based algorithms can distinguish an at-risk patient population with excellent sensitivity and specificity by simply walking, standing, or jumping. This understanding provides a technology that could be deployed in a clinical exam room, athletic training room, strength and conditioning facility, or any number of military settings.

Future studies should pursue the development of MDR-based algorithms to identify those at risk for ACLI before primary injury occurs. This would need to be done through research aimed at completing prospective screening programs where participants perform activities in the presence of MDR. By following the cohort over time and identifying who develops ACLI, algorithms can be developed to identify the highest-risk group, enabling the implementation of validated injury prevention programs. By enabling precise risk stratification, MDR can help target limited preventive resources to those most at risk of injury, in turn, decreasing the total number of participants rather than implementing for the entire population. Other applications could be trained for prognostic and/or diagnostic decision-making in topics such as ankle instability, fall risk, Parkinsons, and cognitive decline.

### 4.3 Limitations

Here, we present findings from a machine learning analysis of MDR data. Although there is a known “black box” phenomenon linked to machine learning, our Threshold-Aware Cascade model confirmed that our model prioritized the STS in classifying the ACLR versus CTRL groups. Based on previous motion capture studies, we can make generalizations regarding biomechanical differences characteristic of ACLR, but future validation would benefit from pairing motion capture comparisons and MDR.

It is worth noting that our study population (n=181), in addition to exhibiting a narrow age range, was homogeneous with regard to race due to the limited geographic region in which the data were collected. Although a younger age range is helpful for active athletes or military applications, more diversity will be needed for future applications involving pediatrics or geriatric populations. We would also note that there was a statistically significant difference in BMI between the ACLR and control groups (ACLR=24.93 and CTRL=26.69), but this difference is so small that it is unlikely to influence our findings. Future studies should control for BMI.

Finally, the predictions here were based on a single variable using only MDR data. MSKIs are complicated and multi-factorial, including intrinsic and extrinsic factors that cannot all be controlled. Future MDR modeling should include comprehensive risk prediction modeling that includes a multivariate risk model with other known risk factors in addition to the MDR prediction.

## 5 Conclusions

There is no current clinically validated efficient approach to objectively evaluate human motion at the point of care. MDR represents a practical technology that has been validated and used in other non-clinical settings for decades. When coupled with machine learning, MDR accurately differentiates ACLR from control groups by identifying complex biomechanical asymmetries, with classification performance comparable to or exceeding that of motion capture. Future research is needed to determine whether MDR can be used in conjunction with risk prediction modeling to identify those most at risk for MSKI as part of injury screening programs.

## Data Availability

All data produced in the present study are available upon reasonable request to the authors

## Acknowledgements

Thank you to the countless family members, mentors, colleagues, support staff, and students who played a role in supporting the acquisition of funding and execution of this research protocol.

## Data Availability

The data are not publicly available because they contain protected health information; access may be granted subject to institutional approval and applicable data use agreements.

## Clinical Trials Registry

ClinicalTrials.gov ID: NCTO5521126

## References

1. Musahl V, Karlsson J. Anterior Cruciate Ligament Tear. N Engl J Med. 2019 Jun 13;380(24):2341–8.

2. Sanders TL, Maradit Kremers H, Bryan AJ, Larson DR, Dahm DL, Levy BA, et al. Incidence of Anterior Cruciate Ligament Tears and Reconstruction: A 21-Year Population-Based Study. Am J Sports Med. 2016 Jun;44(6):1502–7.

3. Dewig DR, Boltz AJ, Moffit RE, Rao N, Collins CL, Chandran A. Epidemiology of Anterior Cruciate Ligament Tears in National Collegiate Athletic Association Athletes: 2014/2015-2018/2019. Med Sci Sports Exerc. 2024 Jan 1;56(1):29–36.

4. Peebles LA, O’Brien LT, Dekker TJ, Kennedy MI, Akamefula R, Provencher MT. The Warrior Athlete Part 2-Return to Duty in the US Military: Advancing ACL Rehabilitation in the Tactical Athlete. Sports Med Arthrosc Rev. 2019 Sep;27(3):e12–e24.

5. Faltstrom A, Kvist J, Bittencourt NFN, Mendonca LD, Hagglund M. Clinical Risk Profile for a Second Anterior Cruciate Ligament Injury in Female Soccer Players After Anterior Cruciate Ligament Reconstruction. Am J Sports Med. 2021 May;49(6):1421–30.

6. Everhart JS, Jones MH, Yalcin S, Reinke EK, Huston LJ, Andrish JT, et al. The Clinical Radiographic Incidence of Posttraumatic Osteoarthritis 10 Years After Anterior Cruciate Ligament Reconstruction: Data From the MOON Nested Cohort. Am J Sports Med. 2021 Apr;49(5):1251–61.

7. Crotti M, Heering T, Lander N, Fox A, Barnett LM, Duncan MJ. Extrinsic Risk Factors for Primary Noncontact Anterior Cruciate Ligament Injury in Adolescents Aged between 14 and 18 years: A Systematic Review. Sports Med. 2024 Apr;54(4):875–94.

8. Kamatsuki Y, Qvale MS, Steffen K, Wangensteen A, Krosshaug T. Anatomic Risk Factors for Initial and Secondary Noncontact Anterior Cruciate Ligament Injury: A Prospective Cohort Study in 880 Female Elite Handball and Soccer Players. Am J Sports Med. 2025 Jan;53(1):123–31.

9. Huang YL, Jung J, Mulligan CMS, Oh J, Norcross MF. A Majority of Anterior Cruciate Ligament Injuries Can Be Prevented by Injury Prevention Programs: A Systematic Review of Randomized Controlled Trials and Cluster-Randomized Controlled Trials With Meta-analysis. Am J Sports Med. 2020 May;48(6):1505–15.

10. Lemes IR, Pinto RZ, Lage VN, Roch BAB, Verhagen E, Bolling C, et al. Do exercise-based prevention programmes reduce non-contact musculoskeletal injuries in football (soccer)? A systematic review and meta-analysis with 13 355 athletes and more than 1 million exposure hours. Br J Sports Med. 2021 May 17;epub 5/19/2021.

11. Thorborg K, Krommes KK, Esteve E, Clausen MB, Bartels EM, Rathleff MS. Effect of specific exercise-based football injury prevention programmes on the overall injury rate in football: a systematic review and meta-analysis of the FIFA 11 and 11+ programmes. Br J Sports Med. 2017 Apr;51(7):562–71.

12. Ross AG, Donaldson A, Poulos RG. Nationwide sports injury prevention strategies: A scoping review. Scand J Med Sci Sports. 2021 Feb;31(2):246–64.

13. Centers for Disease Control and Prevention. COVID-19 Vaccination: Clinical Resources for Each COVID-19 Vaccine. Accessed 8/23/2021 at https://www.cdc.gov/vaccines/covid-19/index.html.

14. U.S. Preventive Services Task Force. USPSTF A and B Recommendations. Accessed 8/23/2021 at https://www.uspreventiveservicestaskforce.org/uspstf/recommendation-topics/uspstf-and-b-recommendations.

15. Chan MS, Sigward SM. Loading Behaviors Do Not Match Loading Abilities Postanterior Cruciate Ligament Reconstruction. Med Sci Sports Exerc. 2019 Aug;51(8):1626–34.

16. Chan MS, Sigward SM. Individuals following anterior cruciate ligament reconstruction practice underloading strategies during daily activity. Journal of Orthopaedic Research®. 2021.

17. Laudani L, Giombini A, Mariani PP, Pigozzi F, Macaluso A. Application of the sit-to-stand movement for the early assessment of functional deficits in patients who underwent anterior cruciate ligament reconstruction. Am J Phys Med Rehabil. 2014 Mar;93(3):189–99.

18. Cabarkapa D, Cabarkapa DV, Song Y, Fry AC, Gisladottir T, Petrovic M. Drop jump performance differences between ACL-injured and healthy semi-professional male soccer players. Front Sports Act Living. 2025;7:1618284.

19. Goerger BM, Marshall SW, Beutler AI, Blackburn JT, Wilckens JH, Padua DA. Anterior cruciate ligament injury alters preinjury lower extremity biomechanics in the injured and uninjured leg: the JUMP-ACL study. Br J Sports Med. 2015 Feb;49(3):188–95.

20. Tsarbou C. Unfolding the complexity of anterior cruciate ligament injury through systems thinking methods (PhD Academy Award). Br J Sports Med. 2025 Sep 2;59(18):1317–8.

21. Leckey C, van Dyk N, Doherty C, Lawlor A, Delahunt E. Machine learning approaches to injury risk prediction in sport: a scoping review with evidence synthesis. Br J Sports Med. 2025 Mar 25;59(7):491–500.

22. Jauhiainen S, Kauppi JP, Krosshaug T, Bahr R, Bartsch J, Äyrämö S. Predicting ACL Injury Using Machine Learning on Data From an Extensive Screening Test Battery of 880 Female Elite Athletes. Am J Sports Med. 2022 Sep;50(11):2917–24.

23. Schweizer N, Strutzenberger G, Franchi MV, Farshad M, Scherr J, Spörri J. Screening Tests for Assessing Athletes at Risk of ACL Injury or Reinjury-A Scoping Review. Int J Environ Res Public Health. 2022 Mar 1;19(5).

24. Bullock GS, Mylott J, Hughes T, Nicholson KF, Riley RD, Collins GS. Just How Confident Can We Be in Predicting Sports Injuries? A Systematic Review of the Methodological Conduct and Performance of Existing Musculoskeletal Injury Prediction Models in Sport. Sports Med. 2022 Oct;52(10):2469–82.

25. Seifert AK, Grimmer M, Zoubir AM. Doppler Radar for the Extraction of Biomechanical Parameters in Gait Analysis. IEEE J Biomed Health Inform. 2021 Feb;25(2):547–58.

26. Imamura RT. Radar as a Means to Study the Biomechanics of Gait: A Validity Study: Georgia State University, Atlanta Ga; 2002.

27. van Dorp P, Groen FCA. Human Walking Estimation with Radar. 2003: IEEE Proc - Radar Sonar Navig; 2003. p. 356–65.

28. Kim M BS, Cha S, Hyun E JY, Bae J, Choi I. Biomechanical Parameters Estimation for Real-Time Gait Analysis Using a Compact Radar Sensor. IEEE Sens J. 2025;25:6620–33.

29. Skolnik MI. Introduction to Radar. New York: Mc-Graw Hill; 1970.

30. Chen VC, Li F, Ho S-S, Wechsler H. Micro-Doppler Effect in Radar: Phenomenon, Model, and Simulation Study. IEEE Trans Aerosp Electron Syst. 2006;42:2–21.

31. Kim Y, Ling H. Human Activity Classification Based on Micro-Doppler Signatures Using an Artificial Neural Network. IEEE Antennas and Propagation Society International Symposium; 2008; San Diego, CA; 2008. p. 1–4.

32. Amin MG, Zhang YD, Ahmad F, Ho KCD. Radar Signal Processing for Elderly Fall Detection: The Future for In-Home Monitoring. IEEE Signal Processing Magazine. 2016:71–80.

33. Narayanan RM, Zenaldin M. Radar micro-Doppler signatures of various human activities. IET Radar, Sonar & Navigation. 2015;9(9):1205–15.

34. Onks C, Hall D, Ridder T, Idriss Z, Andrie J, Narayanan R. The accuracy and predictability of micro Doppler radar signature projection algorithm measuring functional movement in NCAA athletes. Gait Posture. 2021 Mar;85:96–102.

35. Zeng C, Narayanan RM, Onks C. Extending Radar Micro-Doppler Analysis to Various Types of Gait Abnormalities. In: Abagail S. Hedden GJM, editor. SPIE Radar Sensor Technology XXIX Conference; 2025; Orlando, FL: SPIE; 2025.

36. Hayashi S, Saho K, Shioiri K, Fujimoto M, Masugi M. Utilization of Micro-Doppler Radar to Classify Gait Patterns of Young and Elderly Adults: An Approach Using a Long Short-Term Memory Network. Sensors (Basel). 2021 May 24;21(11).

37. Ferdous J, Ahmad F, Onks C. Discriminant analysis of radar micro-Doppler signatures for musculoskeletal injury risk assessment: SPIE; 2025.

38. Ayena JC, Chioukh L, Otis MJ, Deslandes D. Risk of Falling in a Timed Up and Go Test Using an UWB Radar and an Instrumented Insole. Sensors (Basel). 2021 Jan 21;21(3).

39. Abou Al Ardat B, Nyland J, Creath R, Murphy T, Narayanan R, Onks C. Micro-doppler radar to evaluate risk for musculoskeletal injury: Protocol for a case-control study with gold standard comparison. PLoS One. 2023;18(10):e0292675.

40. Harris PA, Taylor R, Thielke R, Payne J, Gonzalez N, Conde JG. Research electronic data capture (REDCap)—a metadata-driven methodology and workflow process for providing translational research informatics support. J Biomed Inform. 2009;42(2):377–81.

41. Hewett TE, Myer GD, Ford KR, Heidt RS, Jr., Colosimo AJ, McLean SG, et al. Biomechanical measures of neuromuscular control and valgus loading of the knee predict anterior cruciate ligament injury risk in female athletes: a prospective study. Am J Sports Med. 2005 Apr;33(4):492–501.

42. Ford KR, Myer GD, Hewett TE. Reliability of landing 3D motion analysis: implications for longitudinal analyses. Med Sci Sports Exerc. 2007 Nov;39(11):2021–8.

43. Ford KR, Myer GD, Hewett TE. Valgus knee motion during landing in high school female and male basketball players. Med Sci Sports Exerc. 2003 Oct;35(10):1745–50.

44. Paterno MV, Schmitt LC, Ford KR, Rauh MJ, Myer GD, Huang B, et al. Biomechanical measures during landing and postural stability predict second anterior cruciate ligament injury after anterior cruciate ligament reconstruction and return to sport. Am J Sports Med. 2010 Oct;38(10):1968–78.

45. Paterno MV, Ford KR, Myer GD, Heyl R, Hewett TE. Limb asymmetries in landing and jumping 2 years following anterior cruciate ligament reconstruction. Clin J Sport Med. 2007;17(4):258–62.

46. Quatman CE, Hewett TE. The anterior cruciate ligament injury controversy: is “valgus collapse” a sex-specific mechanism? Br J Sports Med. 2009 May;43(5):328–35.

47. Kember LS, Riehm CD, Schille A, Slaton JA, Oliver JL, Myer GD, et al. Kinetics During the Tuck Jump Assessment and Biomechanical Deficits in Female Athletes 12 Months After ACLR Surgery. Am J Sports Med. 2025 Feb;53(2):333–42.

48. Chan MS, Sigward SM. Individuals following anterior cruciate ligament reconstruction practice underloading strategies during daily activity. J Orthop Res. 2021 Apr 29;epub 4/30/2021.

49. Capin JJ, Zarzycki R, Arundale A, Cummer K, Snyder-Mackler L. Report of the Primary Outcomes for Gait Mechanics in Men of the ACL-SPORTS Trial: Secondary Prevention With and Without Perturbation Training Does Not Restore Gait Symmetry in Men 1 or 2 Years After ACL Reconstruction. Clin Orthop Relat Res. 2017 Oct;475(10):2513–22.

50. Dewig DR, Johnston CD, Pietrosimone B, Blackburn JT. Long-term gait biomechanics in level, uphill, and downhill conditions following anterior cruciate ligament reconstruction. Clin Biomech (Bristol, Avon). 2021 Apr;84:105345.

51. Zeng C, Narayanan RM, Onks C. Electromagnetic modeling and analysis of micro-Doppler characteristics of simulated human limping gaits. In: Hedden A MG, editor. SPIE. Baltimore; 2024.

52. Alanazi MA, Alhazmi AK, Alsattam O, Gnau K, Brown M, Thiel S, et al. Towards a Low-Cost Solution for Gait Analysis Using Millimeter Wave Sensor and Machine Learning. Sensors (Basel). 2022 Jul 22;22(15).

53. Hall DL, Ridder TD, Narayanan RM. Abnormal gait detection and classification using micro-Doppler radar signatures. Radar Sensor Technology XXIII; 2019: International Society for Optics and Photonics; 2019. p. 110030Q.

54. Alarifi SM, Herrington LC, Althomali OW, Alenezi F, Bin Sheeha B, Jones RK. Biomechanical Analysis After Anterior Cruciate Ligament Reconstruction at the Return-to-Sport Time Point. Orthop J Sports Med. 2025 May;13(5):23259671251340302.

55. Paterno M, Schmitt L, Ford K, Rauh M, Myer G, Huang B, et al. Biomechanical Measures During Landing and Postural Stability Predict Second Anterior Cruciate Ligament Injury After Anterior Cruciate Ligament Reconstruction and Return to Sport. The American journal of sports medicine. 2010 10/01;38:1968–78.

56. Liu L, Popescu M, Skubic M, Rantz M, Yardibi T, Cuddihy P. Automatic fall detection based on Doppler radar motion signature. 2011 5th International Conference on Pervasive Computing Technologies for Healthcare (PervasiveHealth) and Workshops; 2011: IEEE; 2011. p. 222–5.

57. Soubra R, Mourad-Chehade F, Chkeir A. Automation of the Timed Up and Go Test Using a Doppler Radar System for Gait and Balance Analysis in Elderly People. J Healthc Eng. 2023;2023:2016262.

58. Rantz M, Skubic M, Abbott C, Galambos C, Popescu M, Keller J, et al. Automated In-Home Fall Risk Assessment and Detection Sensor System for Elders. Gerontologist. 2015 Jun;55 Suppl 1(Suppl 1):S78–87.

